# The association between at-home exercise digital metrics and ALS disease progression in lower limbs

**DOI:** 10.64898/2026.08.21.26361013

**Authors:** Marcin Straczkiewicz, Narghes Calcagno, Katherine M. Burke, Sravan Mandepudi, Horacio Sanchez Trigo, Alan Premasiri, Fernando G. Vieira, James D. Berry

## Abstract

**Background:** Clinical assessments of Amyotrophic Lateral Sclerosis (ALS) are typically collected infrequently in clinic visits and may not fully capture domain-specific functional decline in daily life. Digital Health Technologies (DHTs) can support remote monitoring, but passive free-living measures often require prolonged wear time and may be influenced by non-motor factors. This study evaluated whether short, standardized, at-home lower limb exercises recorded with ankle-worn accelerometers provide objective and interpretable measures of lower limb disease progression in ALS.

**Methods:** We analyzed data from 349 participants with ALS enrolled in the decentralized ALS Research Collaborative Study. Participants completed repeated self-entry ALS Functional Rating Scale-Revised (ALSFRS-RSE) assessments and wore bilateral ankle accelerometers during monitoring periods between September 2014 and January 2023. During each period, participants performed brief seated knee flexion-extension exercises at home. A previously developed signal processing pipeline was used to derive four exercise metrics: count, duration, intensity, and similarity. We examined baseline correlations with ALSFRS-RSE total and subdomain scores, longitudinal change using linear mixed-effects models, associations with gross motor item scores, differences by anatomical site of disease onset, and comparisons with free-living gait metrics.

**Results:** At baseline, exercise-derived metrics, particularly intensity and similarity, showed the strongest associations with the gross motor subdomain. Longitudinally, duration increased while intensity and similarity decreased, consistent with progressive slowing, reduced movement vigor, and reduced movement consistency (all *p* < 0.001); count did not change significantly. Worsening responses to gross motor items related to turning in bed, walking, and stair climbing were consistently associated with fewer, slower, less vigorous, and less consistent lower limb repetitions. Baseline intensity and similarity were lower in participants with lower limb disease onset on the corresponding side. Exercise-derived intensity showed model fit comparable to the strongest free-living gait metrics, while requiring substantially less observation time.

**Conclusions:** Short at-home lower limb exercises recorded using ankle-worn accelerometers provide scalable, objective, and interpretable measures of amyotrophic lateral sclerosis-related functional decline. Movement quality metrics, particularly intensity and similarity, may complement passive free-living monitoring and support remote digital clinical outcome assessment in ALS research.

**Trial registration:** NCT06885918.

## 1. Introduction

Amyotrophic lateral sclerosis (ALS) is a progressive neurodegenerative disease characterized by motor neuron loss, muscle weakness, and declining ability to perform daily activities ^1,2^. Clinical trials and observational studies in ALS have traditionally relied on clinical outcome assessments such as the ALS Functional Rating Scale-Revised (ALSFRS-R; 12 questions rated 0-4 each with higher scores denoting better function), quantitative muscle strength, and respiratory function. Although these assessments are well established, they are typically collected in clinic at discrete time points and may not fully capture the temporal dynamics or domain-specific manifestations of functional decline ^3,4^. The self-entry ALSFRS-R (ALSFRS-RSE) enables remote assessment, but it remains subjective and ordinal, underscoring the need for objective measures that can be collected at home ^5^.

Digital health technologies (DHTs), including wearable accelerometers and smartphones, offer a scalable approach for measuring human function outside of clinics. By enabling remote and repeated data collection in real-world settings, these tools may reduce the need for frequent clinic visits, decrease participant burden, and support more efficient clinical trials ^6–8^. DHTs can capture both passive data collected continuously or near-continuously during daily activities with minimal user input and active data collected during structured tasks such as surveys, voice recordings, or prescribed exercises ^9^. When used in combination, these passive and active data streams can provide complementary information regarding real-world functional status and longitudinal disease progression ^4,10^.

Despite their potential, important challenges remain. Passive monitoring in free-living settings can capture natural behavior, but it often requires prolonged device wear and sustained participant adherence ^11–13^. In longitudinal studies, adherence may decline over time, and non-wear periods can introduce missing data that complicates analyses and interpretation ^14^. In contrast, traditional in-clinic assessments provide standardized observations but are intermittent, resource intensive, and burdensome for people with progressive neurological disease ^15^. These limitations highlight the need for intermediate approaches that combine the standardization of clinic-based testing with the convenience and scalability of home-based data collection ^16^.

Prescribed exercises performed at home while wearing accelerometers may provide such an intermediate strategy ^10^. In this approach, participants complete short, standardized movement tasks at home while wearing a sensor, allowing quantification of specific motor patterns under more controlled conditions than free-living monitoring. These tasks require substantially less observation time than passive monitoring but may still capture clinically meaningful aspects of motor function capacity. We previously developed an interpretable signal processing method to quantify exercise repetitions from wrist-worn accelerometer data and applied it to upper limb exercises in people living with ALS ^17^. In that work, brief at-home upper limb exercises were used to estimate repetition count, duration, intensity, and similarity, demonstrating that structured movement tasks can sensitively capture upper limb functional decline in ALS.

In the present study, we extended the prescribed exercise framework to assess lower limb function in ALS. Our aim was to establish the clinical validity of wearable sensor-derived metrics of repetition count, duration, intensity, and movement similarity captured during lower limb movements through comparisons with traditional measures, including the ALSFRS-R, as well as with free-living gait metrics captured from passive data collection.

## 2. Methods

### 2.1. Collected data

The data used in this analysis were collected by ALS Therapy Development Institute (ALS TDI) as part of the ALS Research Collaborative Study (ARC), a decentralized observational study of individuals living with ALS, including self-reported ALSFRS-RSE surveys, accelerometer measurements, digital physiological data, speech recordings, and biological samples ^4^.

In the ARC study, clinical function was assessed every 4-6 weeks using the ALSFRS-RSE. The main analyses considered the total ALSFRS-RSE score (Q1—12), subdomain scores for bulbar (Q1—3), fine motor (Q4—6), gross motor (Q7—9), and respiratory (Q10—12) function, and individual gross motor items related to turning in bed (Q7), walking (Q8), and climbing stairs (Q9). The ALSFRS-RSE total score ranges from 0 to 48, while each subdomain ranges from 0 to 12, with higher scores indicating better function. Self-reported anatomical site of disease onset was also used to classify participants according to lower limb onset, with left leg or left foot onset treated as left lower limb onset and right leg or right foot onset treated as right lower limb onset.

Participants wore ActiGraph GT3X+ accelerometers (Ametris, Florida, FL) on both ankles. These devices collected continuous triaxial accelerometer measurements at a sampling frequency of 30 Hz with a dynamic range of ±6 g. Devices were mailed to participants, who were instructed to wear them as much as possible during a 7-11 day monitoring period every 3-4 weeks. During each collection period, participants were asked to complete short lower limb exercise sessions at home and annotate the start and end time of each exercise. The prescribed exercise consisted of repeated seated limb swings, where they were asked to extend their knee to lift their foot off the ground and then lower the foot back down by flexing the knee. They repeated this movements on one side for approximately 45-second period, and then the other side after a 45-second rest period. A YouTube video was provided to guide participants through the session ^18^. In this analysis, we focused on participants who competed prescribed lower limb exercise sessions and repeated ALSFRS-RSE assessments between September 2014 and January 2023.

### 2.2. Digital metrics

We quantified lower limb exercise performance from raw ankle-worn accelerometer data using our previously described signal processing pipeline designed to identify and characterize repeated limb swings during short prescribed exercise tasks ^17^. Raw triaxial acceleration was first transformed into vector magnitude to reduce sensitivity to sensor orientation. Exercise periods were then identified from self-annotated exercise intervals using 1-second non-overlapping windows where the maximum normalized (z-score) signal amplitude exceeded 0.5 SD for at least 10 consecutive seconds. The signal was then decomposed into its time-frequency representation using continuous wavelet transform. The resulting time-frequency decomposition was split into 0.5-second windows, and a comb function was used to identify the fundamental swing frequency and its higher harmonics. The estimated swing frequency was then used to segment the signal into individual limb swings, which were rescaled and time-aligned to enable comparison of movement patterns across repetitions.

From the segmented exercise data, we derived four digital metrics describing complementary aspects of lower limb exercise performance: count (reflecting quantity), duration (speed), intensity (vigor), and similarity (consistency). Count was calculated as the total number of identified limb swings, with correction for the 0.5-second windows used in the time-frequency decomposition. Duration was calculated as the mean inverse of the estimated limb swing frequencies. Intensity was calculated as the mean acceleration of segmented and time-scaled limb swing time series, while similarity was calculated as the mean pairwise correlation between segmented, rescaled, and time-aligned limb swing repetitions. Duration was expressed in seconds, intensity was expressed in gravitational units (g’s), while similarity was unitless.

For comparison with exercise-derived metrics, we also extracted free-living gait metrics using a previously developed approach ^19,20^. These free-living metrics included step counts, top two-minute cadence, stride intensity, stride similarity, stride variability, and stride fragmentation. Free-living gait analyses were restricted to data collected during days of exercise with at least 16 hours of wear-time.

### 2.3. Statistical analysis

The analytic dataset consisted of lower limb exercise metrics and ALSFRS-RSE survey responses from participants who had at least two at-home bilateral exercise sessions and corresponding ALSFRS-RSE assessments completed within 14 days from the first or last exercise day. We summarized participant characteristics, survey completion, and exercise availability using mean and standard deviation (SD) for continuous variables and counts and percentages for categorical variables. Exercise metrics were derived separately for each ankle. The combined metrics considered lower exercise count, intensity, and similarity from either ankle, and longer duration from either ankle. The main analysis focused on metrics from the left ankle, while corresponding analyses for the right ankle and bilateral metrics are reported in the **Supplementary Materials**.

For baseline analyses, we used the first valid set of exercise-derived metrics and the corresponding baseline ALSFRS-RSE survey. Pearson correlation coefficients were calculated between the four exercise metrics, i.e., count, duration, intensity, and similarity, and ALSFRS-RSE total and subdomain scores.

To quantify longitudinal change, we fit linear mixed-effect models (LMMs) with participant-specific random intercepts and random slopes for time. In these models, time elapsed from enrollment was included as a fixed effect and expressed in months, enabling estimation of baseline values and monthly rates of change for each exercise metric and ALSFRS-RSE score. These models were used to assess whether lower limb exercise metrics changed over time and whether their longitudinal patterns were consistent with progressive functional decline measured by ALSFRS-RSE. We reported baseline estimates, monthly change estimates, 95% confidence intervals, *p*-values, conditional and marginal R-squared values, standardized annual change (in SD units), and relative annual change (%).

We also fit LMMs to assess associations between exercise-derived metrics and individual ALSFRS-RSE items related to lower limb and gross motor function (one model per question). Specifically, we examined Q7 (turning in bed and adjusting bed clothes), Q8 (walking ability), and Q9 (stair climbing). In these models, each exercise metric was modeled as the outcome and the ALSFRS-RSE item response was modeled as a categorical predictor. A response equal to 4, corresponding to normal function, was treated as the reference category. This approach allowed us to estimate how each level of self-reported impairment was associated with count, duration, intensity, and similarity of at-home lower limb exercise.

To compare short prescribed exercises with free-living movement monitoring, we then fit an additional LMMs to assess associations between ALSFRS-RSE total score and both exercise-derived and free-living gait metrics, and compared their model fit and association with overall functional status.

Finally, we evaluated whether baseline digital exercise metrics differ according to self-reported anatomical site of disease onset and baseline gait performance. For onset analyses, left leg or left foot onset was classified as left lower limb onset, while right leg or right foot onset was classified as right lower limb onset. Two-sample t-tests were used to compare baseline exercise metrics between participants with and without onset on the corresponding lower limb. For gait performance stratification, participants who scored 4 on both ALSFRS-RSE Q8 and Q9 at baseline were classified as having preserved lower limb function, whereas participants with any score < 4 on either item were classified as having impaired lower limb function. Longitudinal models were then fit separately within these strata, and slope differences were used to compare rates of change between participants for each group.

## 3. Results

### 3.1. Population

Out of 389 participants with accelerometer data, 32 did not contribute longitudinal data, 6 had data from a single ankle only, and 2 had no corresponding survey responses. The analytic cohort of 349 participants with ALS (**Table 1**) predominantly involved white (95.42%) males (65.33%) with a mean (SD) age of 55.33 (10.69) years and a mean (SD) body mass index (BMI) of 25.97 (4.65) kg/m². Participants completed a mean (SD) of 30.79 (26.15) ALSFRS-RSE surveys, with an average (SD) interval of 38.31 (26.97) days between surveys. The average single-limb exercise duration closely matched the prescribed 45-second protocol, with mean (SD) durations of 46.39 (1.70) seconds for the left ankle and 45.55 (1.91) seconds for the right.

**Table 1.**
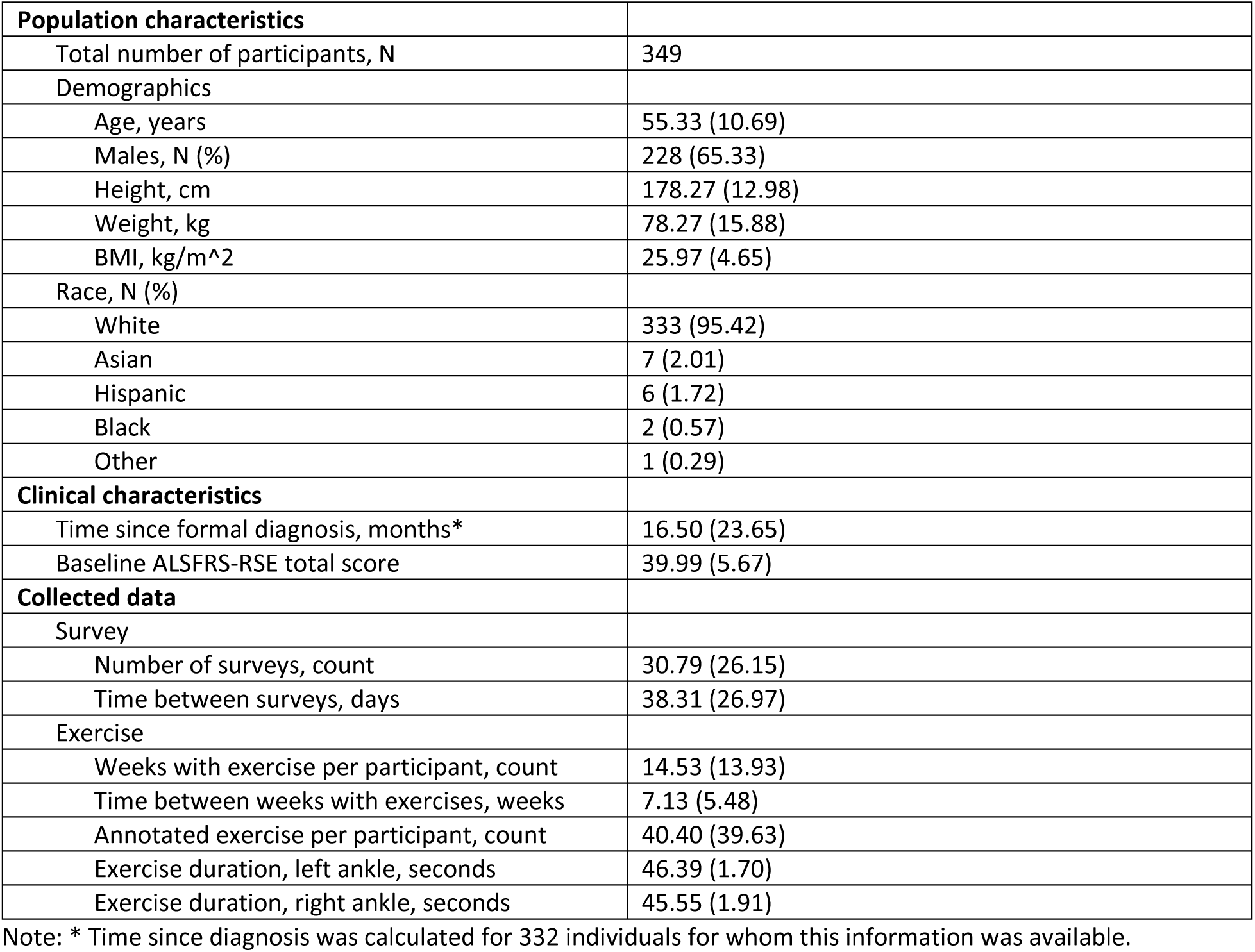
Summary on population and clinical characteristics, collected surveys, and exercise statistics. Mean (SD) unless indicated otherwise.

### 3.2. Baseline correlation between digital metrics and ALSFRS-RSE

At baseline, the four exercise-derived lower limb swing metrics demonstrated expected interrelationships (**Figure 1**). Count was strongly negatively correlated with duration (Pearson r = −0.78) and moderately positively correlated with intensity (r = 0.57), indicating that participants who performed more repetitions generally performed them faster and with greater movement intensity. Duration was negatively correlated with intensity (r = −0.59) and similarity (r = −0.44), while intensity was moderately correlated with similarity (r = 0.54). These findings indicate that the derived metrics captured related but distinct aspects of lower limb exercise performance, including movement quantity, speed, vigor, and consistency.

**Figure 1.**
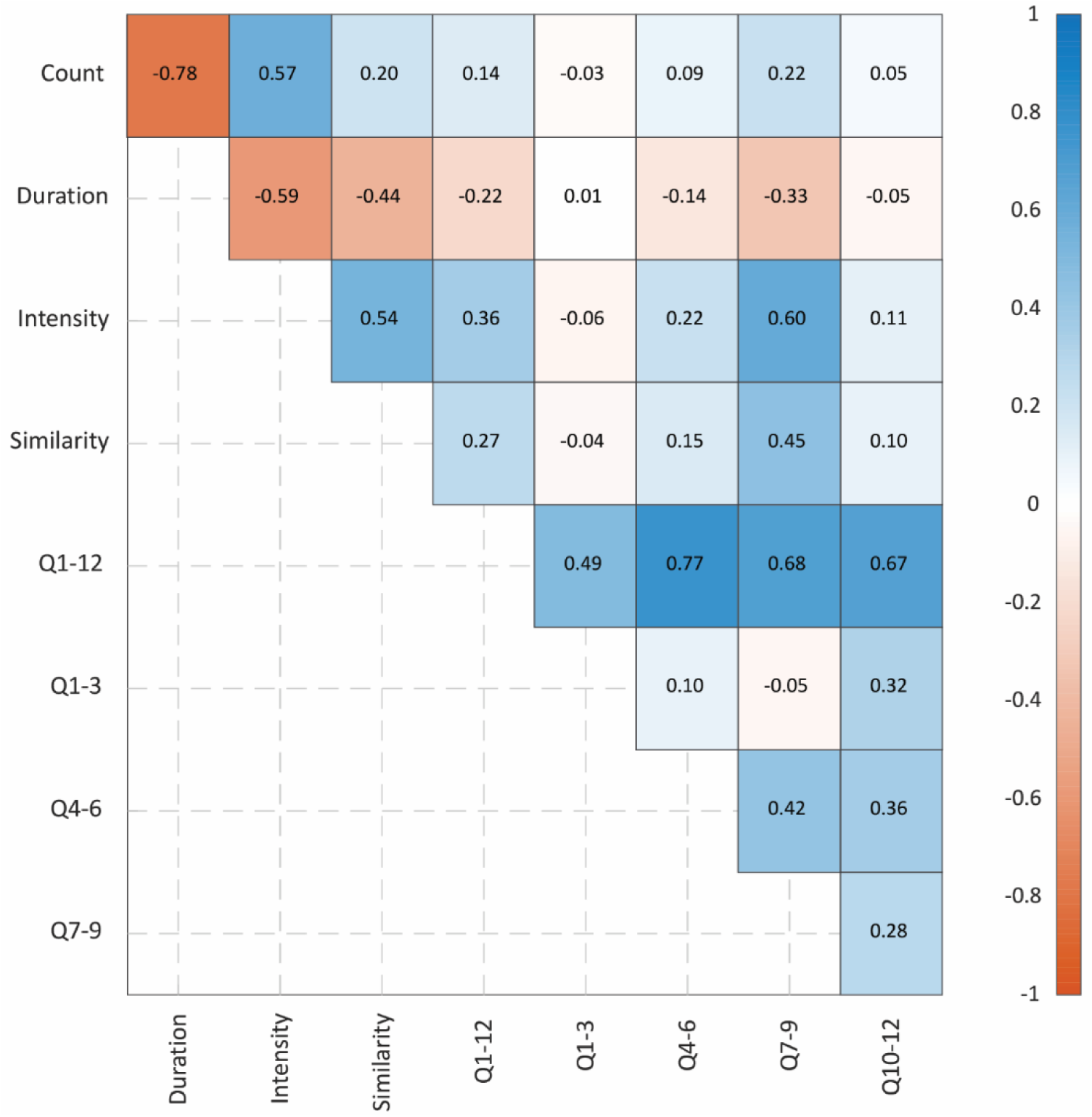
Pearson correlation coefficients. Correlation coefficients were estimated between baseline limb swing measures and ALSFRS-RSE total score (Q1—12) and subdomain scores (Q1—3 bulbar, Q4—6 fine motor, Q7—9 gross motor, Q10—12 respiratory).

Correlations between exercise-derived metrics and ALSFRS-RSE scores were weaker than the correlations among the digital metrics themselves. The ALSFRS-RSE total score (Q1—12) showed weak positive correlations with count (r = 0.14), intensity (r = 0.36), and similarity (r = 0.27), and a weak negative correlation with duration (r = −0.22; movements were performed faster when ALSFRS-R scores were higher). Among ALSFRS-RSE subdomains, the strongest associations were observed for gross motor function (Q7—9), with correlations of r = 0.22 for count, r = −0.33 for duration, r = 0.60 for intensity, and r = 0.45 for similarity. In contrast, correlations with bulbar (Q1—3) and respiratory (Q10—12) subdomains were negligible to weak, ranging from −0.06 to 0.11 for the former and from −0.05 to 0.11 for the latter across the four exercise metrics. Overall, the strongest baseline correlations with ALSFRS-RSE were observed for intensity and similarity in relation to gross motor function, consistent with the intended lower limb focus of the exercise task.

The supplementary baseline analysis of the digital metrics from the right and both ankles were consistent with the findings from the primary left ankle analysis (**Figure S1**).

### 3.3. Longitudinal change

Using LMMs, we determined that three out of four left ankle exercise metrics changed significantly over time (**Table 2**). Specifically, limb swing duration increased, while intensity and similarity decreased (all *p* < 0.001). Limb swing count did not demonstrate statistically significant change (*p* = 0.081). These results indicate that, while participants maintained a similar number of repetitions over time, their movements became slower, less vigorous, and less consistent.

**Table 2.** Average baseline and monthly change in the left limb swing movements metrics and ALSFRS-RSE scores. Standardized and relative changes are expressed as SD or % per 52 weeks, respectively.

| Outcome | Baseline estimate [95% CI] | Monthly change estimate [95% CI] | p-value | R2c | R2m | Standardized change (SD per year) | Relative change (% per year) |
| --- | --- | --- | --- | --- | --- | --- | --- |
| Count | 45.11 [43.96, 46.25] | -0.072 [-0.153, 0.009] | 0.081 | 0.869 | 0.005 | -0.060 | -1.92 |
| Duration | 1.159 [1.122, 1.195] | 0.007 [0.003, 0.011] | <0.001 | 0.885 | 0.022 | 0.165 | 7.38 |
| Intensity | 0.550 [0.515, 0.584] | -0.007 [-0.009, -0.005] | <0.001 | 0.935 | 0.059 | -0.205 | -14.72 |
| Similarity | 0.854 [0.837, 0.871] | -0.002 [-0.003, -0.001] | <0.001 | 0.850 | 0.021 | -0.135 | -2.70 |
| Q1—3 | 10.51 [10.28, 10.74] | -0.118 [-0.138, -0.097] | <0.001 | 0.991 | 0.132 | -0.517 | -13.68 |
| Q4—6 | 9.36 [9.09, 9.64] | -0.168 [-0.188, -0.147] | <0.001 | 0.988 | 0.264 | -0.692 | -21.94 |
| Q7—9 | 9.13 [8.85, 9.40] | -0.192 [-0.213, -0.171] | <0.001 | 0.988 | 0.321 | -0.810 | -25.69 |
| Q10—12 | 11.20 [11.03, 11.37] | -0.070 [-0.084, -0.056] | <0.001 | 0.975 | 0.125 | -0.419 | -7.64 |
| Q1—12 | 40.24 [39.62, 40.86] | -0.564 [-0.625, -0.503] | <0.001 | 0.994 | 0.322 | -0.900 | -17.16 |

Note: R2c – R-squared conditional, R2m – R-squared marginal.

The ALSFRS-RSE scores also declined significantly over time across all domains. For example, the total ALSFRS-RSE score (Q1—12) decreased by −0.564 points per month, corresponding to a standardized annual change of −0.900 SD and a relative annual change of −17.16%. Among subdomains, the largest standardized annual decline was observed for gross motor function, followed by fine motor function, bulbar function, and respiratory function (all *p* < 0.001). Among the digital exercise metrics, intensity exhibited the largest annual standardized change (−0.205 SD; relative change −14.72%), followed by duration (0.165 SD; relative change 7.38%) and similarity (−0.135 SD; relative change −2.70%). Thus, longitudinal changes in exercise quality metrics were directionally consistent with ALS disease progression, with intensity showing the most pronounced decline among the digital measures.

The sensitivity analysis (**Supplementary Materials, Table S1**) showed that the longitudinal patterns observed for the left ankle were reproduced when exercise metrics were derived from the right ankle and from both ankles combined.

### 3.4. Digital metrics vs. ALSFRS-RSE

We fit LMMs to assess associations between left ankle exercise metrics and ALSFRS-RSE items related to gross motor function (Q7—9), with normal function (score = 4) used as the reference category for each item.

Compared with participants reporting normal function on Q7 (turning in bed and adjusting the bed clothes), those with lower Q7 scores completed fewer repetitions, at slower speeds, lower intensities, and with lower consistency (**Figure 2A**). These differences became progressively more pronounced as Q7 scores decreased. All Q7 associations were statistically significant, with *p* < 0.001 except for count at Q7 = 3 (*p* = 0.014) and duration at Q7 = 0 (*p* = 0.007). Similar patterns were seen for both Q8 (walking, **Figure 2B**) and Q9 (climbing stairs, **Figure 2C**). Most associations were statistically significant for Q8; the only exception was count for Q8 = 3, where the confidence interval included zero (−1.29; 95% CI −2.65, 0.07; *p* = 0.063). All duration, intensity, and similarity associations were statistically significant at *p* < 0.001. For count, associations were significant for Q9 = 2, Q9 = 1, and Q9 = 0, but not for Q9 = 3 (−0.92; 95% CI −2.24, 0.39; *p* = 0.167). Overall, worsening responses to gross motor ALSFRS-RSE items were consistently associated with fewer, slower, less vigorous, and less consistent lower limb exercise repetitions.

**Figure 2.**
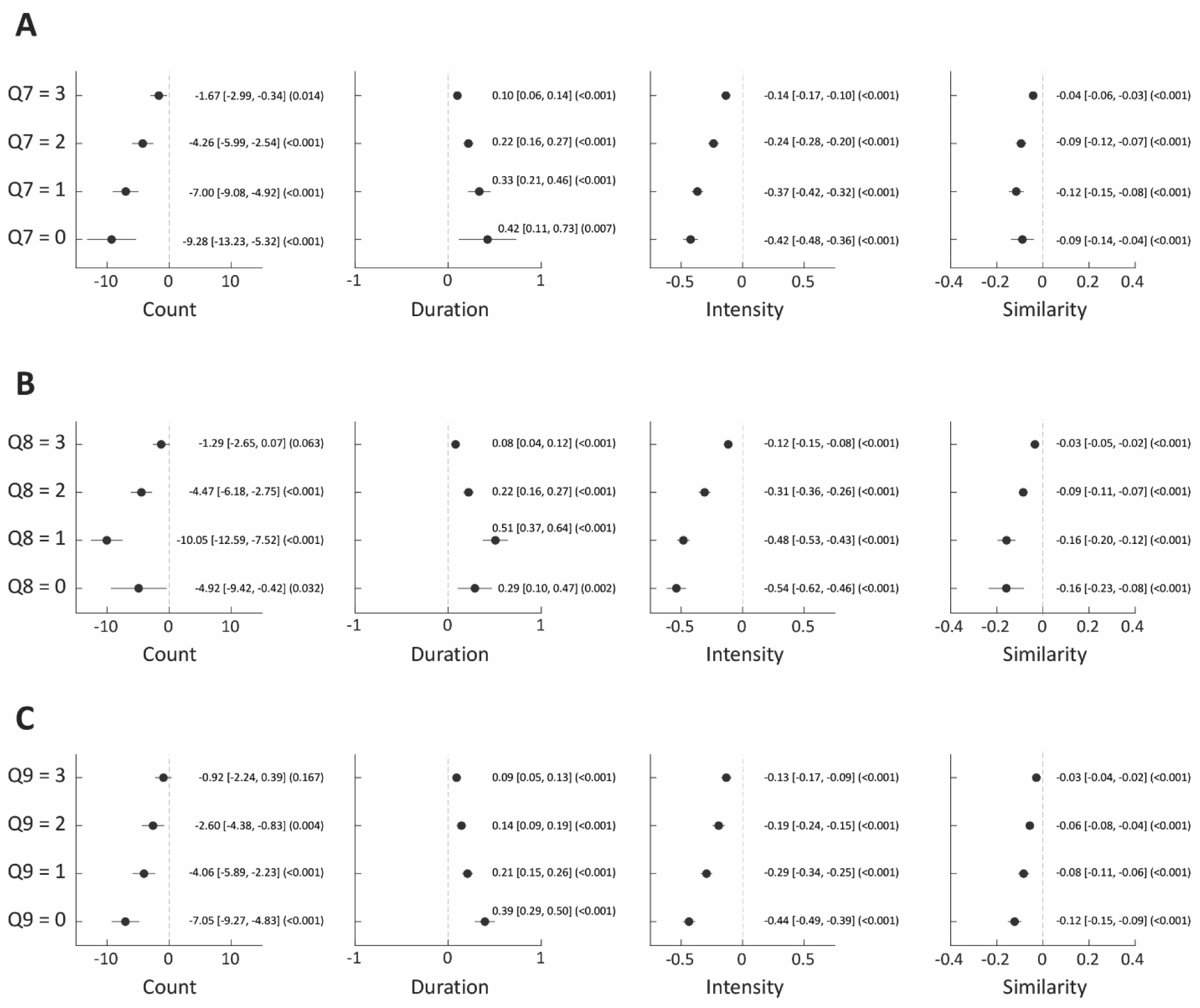
Forest plot of estimated regression coefficients (95% confidence intervals, *p*-value) for ALSFRS-RSE turning in bed (Q7, panel A), walking (Q8, B), and stair climbing (Q9, C). Participants reporting normal functioning (response equal to 4) served as the reference value (dashed line).

The summary of underlying data used for modelling the association between digital exercise metrics and ALSFRS-RSE were provided in **Supplementary Materials** (**Table S2**).

### 3.5. Exercise vs. free-living metrics

We next compared exercise-derived metrics with free-living gait metrics in observations with simultaneous exercise data and substantial sensor wear time (≥16 h) (**Table 3**). After applying this criterion, the analytic sample included 249 participants (28.7% decrease). Among exercise-derived metrics, higher ALSFRS-RSE total score (Q1— 12) was significantly associated with higher count (*p* = 0.016), higher intensity (*p* = 1.18e-12), and higher similarity (*p* = 0.001). Duration showed the expected negative association with Q1—12, but this association did not reach statistical significance (*p* = 0.092).

**Table 3.** Model intercept and estimated average change associated with a one-point increase in ALSFRS-RSE Q1— 12 using limb swing-derived outcomes of accelerometry data for the free-living lower limb movements. Models were estimated using data from days with simultaneous exercise and substantial sensor wear time (>=16 h).

| Data collection | Digital metric | Intercept [95% CI] | Slope [95% CI] | p-value | R2c | R2m |
| --- | --- | --- | --- | --- | --- | --- |
| Exercise |  |  |  |  |  |  |
|  | Count | 36.28 [34.47, 38.09] | 0.044 [0.008, 0.080] | 0.016 | 0.813 | 0.008 |
|  | Duration | 39.42 [37.85, 40.99] | -1.11 [-2.41, 0.183] | 0.092 | 0.829 | 0.005 |
|  | Intensity | 34.73 [33.38, 36.08] | 8.22 [5.96, 10.47] | 1.18e-12 | 0.915 | 0.133 |
|  | Similarity | 34.97 [32.91, 37.04] | 3.60 [1.41, 5.79] | 0.001 | 0.784 | 0.008 |
| Free-living |  |  |  |  |  |  |
|  | Step counts | 35.03 [34, 36.06] | 0.001 [0.001, 0.001] | 1.25e-25 | 0.859 | 0.161 |
|  | Cadence | 33.83 [32.66, 34.99] | 0.064 [0.052, 0.075] | 9.72e-26 | 0.830 | 0.099 |
|  | Intensity | 32.05 [30.61, 33.49] | 7.01 [5.73, 8.29] | 1.73e-26 | 0.856 | 0.143 |
|  | Similarity | 35.3 [33.7, 36.89] | 5.15 [2.74, 7.56] | 2.93e-05 | 0.790 | 0.017 |
|  | Variability | 43.01 [41.47, 44.55] | -11.46 [-14.99, -7.92] | 2.41e-10 | 0.817 | 0.048 |
|  | Fragmentation | 43.3 [42.16, 44.45] | -9.97 [-12.08, -7.88] | 2.48e-20 | 0.828 | 0.084 |

Note: R2c – R-squared conditional, R2m – R-squared marginal.

Free-living gait metrics were also significantly associated with ALSFRS-RSE Q1—12. Higher scores were associated with higher step counts (*p* = 1.25e-25), higher cadence (*p* = 9.72e-26), higher stride intensity (*p* = 1.73e-26), and higher stride similarity (*p* = 2.93e-05). In contrast, higher total score was associated with lower stride variability (*p* = 2.41e-10) and stride fragmentation (*p* = 2.48e-20), consistent with better-preserved gait being characterized by more regular and less fragmented walking patterns. Model fit was comparable across free-living metrics.

In the supplementary longitudinal comparison based on left ankle sensor data, free-living gait metrics showed more pronounced temporal change than exercise-derived metrics in the subset of observations with simultaneous exercise data and substantial wear time (**Table S3**). These findings suggest that free-living gait metrics may be more responsive to longitudinal change when sufficient wear time is available, whereas short exercise-derived metrics provide a complementary, lower-burden assessment with meaningful cross-sectional association to ALSFRS-RSE.

### 3.6. Disease onset vs. baseline digital metrics

We compared baseline left ankle exercise metrics between participants with and without left lower limb ALS (**Figure 3**). Overall, 154 participants reported onset in either lower limb, including 54 with left lower limb onset only, 56 with right lower limb onset only, and 44 with bilateral lower limbs; the remaining 195 participants did not report onset in either lower limb. At baseline, participants with left lower limb onset performed fewer repetitions and showed longer movement duration compared with participants without left lower limb onset, but these differences did not reach statistical significance for count (*p* = 0.141) or duration (*p* = 0.100). In contrast, participants with left lower limb onset had significantly lower exercise intensity (*p* = 4.80e-06) and lower similarity (*p* = 0.001), indicating less vigorous and less consistent lower limb movements on the side of symptom onset. These findings suggest that baseline exercise quality metrics, particularly intensity and similarity, were sensitive to the anatomical site of disease onset, whereas repetition count and duration were less clearly differentiated between onset groups.

**Figure 3.**
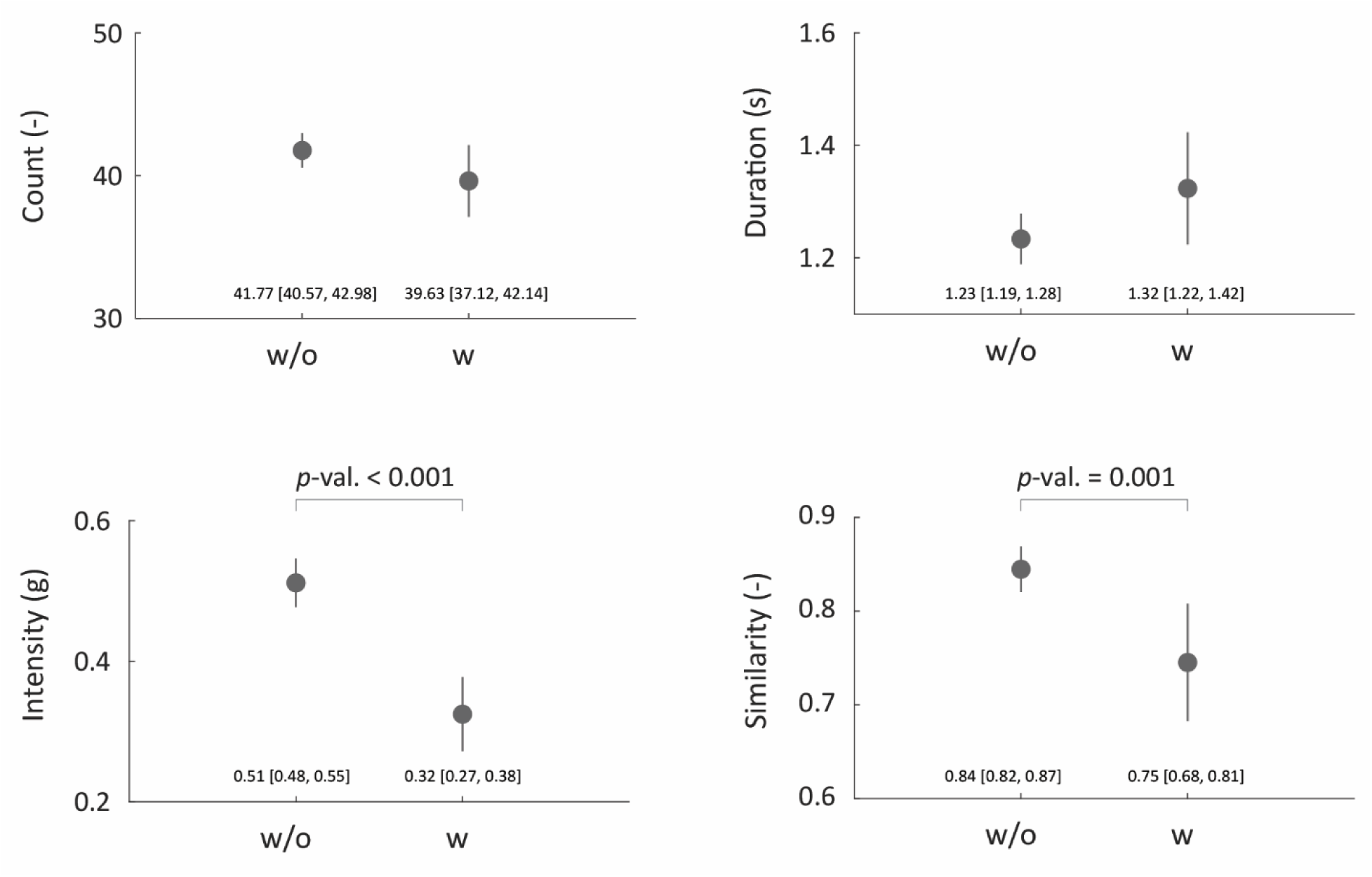
Mean and 95% confidence intervals for baseline digital metrics (first valid set of observations) stratified on participants with (w) and without (w/o) ALS onset on left lower limb.

Similar findings were observed using right ankle metrics (**Supplementary Materials, Figures S2-S3**), while the analysis using metrics derived from both ankles suggest that side-specific sensor placement may be more sensitive to the anatomical site of ALS onset than bilateral aggregation of metrics, particularly for detecting reduced movement intensity and consistency in the affected lower limb.

### 3.7. Stratification based on baseline gait performance

We stratified participants according to baseline lower limb functioning using ALSFRS-RSE Q8 and Q9 (**Table 4**). Participants who reported normal walking (Q8 = 4) and stair climbing (Q9 = 4) at baseline were classified as preserved functioning (n = 100), whereas those with score < 4 on either item were classified as impaired functioning (n = 249). At baseline, the normal-functioning group performed more left limb swing repetitions than those with impaired functioning, with shorter duration, higher intensity, and higher similarity. These baseline differences indicate that lower limb exercise metrics were already separated by baseline self-reported gait status at study entry.

**Table 4.** Comparison between longitudinal changes in left ankle exercise metrics between individuals reporting normal and deteriorated functioning, as reflected on questions Q8 and Q9 in ALSFRS-RSE. Model slopes express monthly change in digital metrics and survey scores. Standardized and relative changes are expressed in SD and percentage change per year, respectively.

| Digital metric | Intercept [95% CI] | Slope [95% CI] | p-value | R2c | R2m | Standardized change (SD per year) | Relative change (% per year) | Intercept [95% CI] | Slope [95% CI] | p-value | R2c | R2m | Standardized change (SD per year) | Relative change (% per year) | Slope difference p-value |
| --- | --- | --- | --- | --- | --- | --- | --- | --- | --- | --- | --- | --- | --- | --- | --- |
|  | Normal at baseline, n=100 |  |  |  |  |  |  | Deteriorated at baseline, n=249 |  |  |  |  |  |  |  |
| Count | 49.97<br>[48.12, 51.82] | 0.013<br>[-0.010, 0.037] | 0.266 | 0.777 | 0.005 | 0.050 | 1.35 | 43.18<br>[41.83, 44.52] | -0.030<br>[-0.055, -0.005] | **0.017** | 0.882 | 0.016 | -0.109 | -3.64 | **0.024** |
| Duration | 0.996<br>[0.960, 1.03] | -0.000<br>[-0.001, 0.000] | 0.255 | 0.866 | 0.007 | -0.084 | -1.97 | 1.22<br>[1.18, 1.27] | 0.002<br>[0.001, 0.004] | **<0.001** | 0.890 | 0.039 | 0.222 | 10.45 | **0.010** |
| Intensity | 0.794<br>[0.741, 0.847] | -0.001<br>[-0.002, -0.000] | **0.024** | 0.840 | 0.028 | -0.152 | -7.70 | 0.451<br>[0.414, 0.487] | -0.002<br>[-0.002, -0.001] | **<0.001** | 0.952 | 0.095 | -0.265 | -19.45 | 0.382 |
| Similarity | 0.926<br>[0.912, 0.939] | -0.000<br>[-0.000, 0.000] | 0.400 | 0.595 | 0.004 | -0.053 | -0.52 | 0.825<br>[0.804, 0.847] | -0.001<br>[-0.001, -0.000] | **<0.001** | 0.862 | 0.032 | -0.164 | -3.74 | 0.051 |

Note: R2c – R-squared conditional, R2m – R-squared marginal.

Longitudinal change also differed by baseline gait performance. Among participants with normal lower limb function at baseline, neither count (*p* = 0.266), nor did duration (*p* = 0.255) or similarity (*p* = 0.400) changed significantly over time. Intensity declined significantly in this group (*p* = 0.024), corresponding to a standardized annual change of −0.152 SD and a relative annual change of −7.70%. In contrast, participants with impaired baseline lower limb functioning showed significant longitudinal changes in all four exercise metrics: count decreased (*p* = 0.017), duration increased (*p* < 0.001), intensity decreased (*p* < 0.001), and similarity decreased (*p* < 0.001).

The between-group comparison of slopes indicated that deterioration over time was more pronounced among participants with impaired baseline gait function for count and duration. Specifically, slope differences were statistically significant for count (*p* = 0.024) and duration (*p* = 0.010), but not for intensity (*p* = 0.382). The difference in similarity slopes approached statistical significance (*p* = 0.051). These results suggest that participants with impaired gait performance at baseline showed significantly more decline in both movement quantity and movement duration, whereas intensity is capturing decline that was similar in both groups, perhaps indicating that intensity can capture early changes.

Results provided in **Supplementary Materials** (**Table S4**) extended this analysis to right ankle and bilateral exercise metrics and were consistent with left-ankle analyses.

## 4. Discussion

In this study, we evaluated whether metrics derived from short, standardized, at-home lower limb exercises recorded using ankle-worn accelerometers can quantify ALS disease progression. We extended our previously developed upper limb exercise quantification framework to lower limb function and derived four interpretable metrics from repeated limb swings: count, duration, intensity, and similarity. In a longitudinal cohort of 349 participants with ALS, these metrics were associated with functional scores captured by the ALSFRS-RSE, changed over time in directions consistent with disease progression, showed good construct criterion validity with the gross motor subdomain of the ALSFRS-RSE, and reflected the anatomical site of lower limb disease onset. We further showed that models fit using prescribed exercise metrics, particularly intensity, were comparable to free-living gait metrics that require substantially longer sensor wear time and that participants are ambulatory. These findings support the use of brief structured exercises as a low-burden, scalable, and interpretable approach to remote lower limb monitoring in ALS.

In this analysis, the strongest and most consistent findings were observed for movement quality rather than the number of repetitions – consistent to our previous findings in the upper limb analysis ^17^. At baseline, count, duration, intensity, and similarity were interrelated in expected ways: participants who completed more repetitions tended to complete them faster and with greater intensity, while longer movement duration was associated with lower intensity and lower similarity. However, in longitudinal models, quality metrics changed more consistently that movement quantity. This could in part reflect the exercise protocol, which instructed participants to perform repetitions in a standardized time period if possible. As a result, count did not significantly change in the primary left ankle analysis, while duration increased and both intensity and similarity decreased over time, capturing movement deterioration despite a relatively stable number of repetitions.

The association between exercise metrics and ALSFRS-RSE scores demonstrates validity and further supports the clinical relevance of the proposed exercise metrics. As expected, baseline correlations were strongest for the gross motor subdomain, especially for intensity and similarity, whereas correlations with bulbar and respiratory subdomains were weak or negligible. This domain specificity is important because a useful digital measure should align most strongly with the functional domain it is intended to quantify. Similarly, in categorical models using individual ALSFRS-RSE items, worse scores on Q7, Q8, and Q9 were consistently associated with fewer, slower, less vigorous, and less consistent lower limb repetitions. These findings suggest that the proposed measures captured aspects of lower limb function relevant not only to walking and stair climbing, but also to broader gross motor tasks such as turning in bed and adjusting bed clothes. This may reflect the shared dependence of these activities on lower limb strength, coordination, and movement control.

Our findings build on prior work demonstrating that DHTs can quantify ALS progression. Johnson et al. showed that wearable devices and smartphone-based self-entry surveys can be collected remotely in people with ALS and that several wearable-derived physical activity measures change over time and associate with ALSFRS-RSE and ROADS ^3^. Karas et al. extended this evidence to passively collected smartphone accelerometer and GPS data, showing that selected smartphone-derived measures of walking and activity changed over time and were associated with ALSFRS-RSE ^21^. In the independent longitudinal cohort, Holdom et al. showed that wrist-worn accelerometer metrics declined over time, correlated with ALSFRS-R motor subdomain, and provided information complimentary to ALSFRS-R, and suggested peak 6-minute activity as a potentially useful outcome measure for long-duration trials ^22^. Other work using wrist-worn accelerometers demonstrated that free-living upper limb movement metrics are associated with ALSFRS-RSE, change longitudinally, and reflect disease onset location ^23^. Lukac et al. focused on lower limb gait using foot-worn inertial sensors and demonstrated that repeated at-home walking speed measurements were feasible in ALS, declined over 24 weeks in most participants, and showed greater decline among those who later transitioned to an assistive device ^24^. The present study adds to this literature by focusing on short standardized active tasks rather than passive monitoring alone, and by applying this framework to lower limb function.

The comparison between the short prescribed exercises and free-living gait metrics is particularly important. Passive free-living monitoring captures natural behavior and can provide rich information about walking volume, cadence, stride intensity, stride similarity, variability, and fragmentation. However, free-living gait metrics require sufficient wear time and depend on whether participants actually walk during the observation window. In ALS, this requirement can become increasingly challenging as mobility declines. In this analysis, exercise-derived intensity achieved model fit comparable to the strongest free-living gait metrics, including stride intensity, step counts, and walking fragmentation. As a result, prescribed exercises may be best viewed as a strategy that provides standardized, interpretable, and analyzable movement data when prolonged wear time or sufficient free-living walking data is difficult to obtain.

Stratification by baseline gait performance also demonstrated that digital exercise metrics captured meaningful differences between participants with preserved and impaired lower limb function. Participants reporting preserved walking and stair climbing at baseline had higher count, higher intensity, higher similarity, and shorter duration than those with impaired lower limb function. Longitudinally, participants with deteriorated baseline function showed significant decline across all four left ankle exercise metrics, while for normal-functioning group, change mainly reflected in declining intensity. Slope differences were significant for count and duration, suggesting that participants with baseline impairment experienced more pronounced changes in movement quantity and speed. These results indicate that prescribed exercise metrics may be informative across disease stages, while also capturing different dimensions of decline depending on baseline functional status.

In a subset of participants with reported onset location, side-specific exercise metrics helped preserve information related to reported anatomical onset location. ALS usually begins focally, spreads locally and then to adjacent body regions ^25,26^. In participants without leg involvement at study baseline, most gait parameters did not change over the observation period. Our findings suggest that side-specific sensors may provide a means of tracking the anatomical spread of the diseases over time. Similar observations have been reported using upper limb assessement^27^. In the future, such digital measures may provide useful for participant inclusion, stratification, subgroup analyses or monitoring regional progression in clinical trials.

Our study did have several limitations. First, exercise timing and task performance were self-annotated by participants in a home setting, and we could not directly verify whether all participants performed the prescribed movements exactly as instructed. Caregiver assistance, incomplete effort, misunderstanding of instructions, or other deviations from the protocol may have influenced the measurements ^28^. Second, while ankle placement is appropriate for lower limb exercise and gait assessment, incorrect device placement or side switching could affect side-specific analyses, particularly those related to disease onset ^29^. Third, the ALSFRS-RSE in an imperfect clinical anchor as it is subjective, ordinal, and multidimensional ^20,30,31^. Fourth, the cohort was predominantly white males with relatively slower rate of progression which may limit generalizability to more diverse populations. Also, participants who contributed data may represent more adherent or less impaired subset of the ARC cohort, while missingness may be related to disease severity, fatigue, mobility impairment, or caregiver support, and should be investigated further. Finally, this study was observational and could not evaluate responsiveness to treatment or establish thresholds for clinically meaningful change.

Future work should validate these metrics in independent cohorts, incorporate direct or automated checks of exercise adherence, and determine the optimal frequency and duration of prescribed lower limb tasks. Studies should also evaluate minimal detectable change and clinically meaningful change using patient- and clinician-centered anchors ^32^. Because gait and lower limb function are closely related to independence, fall risk, and quality of life, future analyses should examine how changes in prescribed exercise metrics map onto outcomes that are directly meaningful to people living with ALS ^28,33^. More broadly, combining prescribed exercise metrics with free-living gait measures, upper limb movement measures, speech, respiratory outcomes, and patient-reported assessments may provide a more comprehensive and domain-specific picture of ALS progression ^34^.

In conclusion, short at-home lower limb exercises recorded with ankle-worn accelerometers provide clinically meaningful, objective, and interpretable measures of ALS-related functional decline. Movement quality metrics, particularly intensity and similarity, were most consistently associated with gross motor function, longitudinal decline, disease onset, and baseline gait impairment. These findings suggest that prescribed exercise assessments can complement passive free-living monitoring and may be especially useful when prolonged sensor wear or free-living walking data are difficult to obtain. By providing standardized assessments with minimal participant burden, this approach may support more scalable remote monitoring and contribute to the development of digital clinical outcome assessments for ALS research and future clinical trials.

## Data Availability

Data may be shared upon request and after review and approval by the owners of the data. Data may be requested through the ALS TDI ARC Data Commons (https://www.als.net/arc/data-commons/).

## Acknowledgements

The authors thank the participants and their caregivers who devoted their time to participate in the study.

## Abbreviations

ALS: Amyotrophic lateral sclerosis
ALSFRS-R: ALS Functional Rating Scale-Revised
ALSFRS-RSE: ALS Functional Rating Scale-Revised Self-Entry
ALS TDI: ALS Therapy Development Institute
ARC: ALS Research Collaborative
BMI: Body mass index
CI: Confidence interval
CIRBI: Center for IRB Intelligence
DHT: Digital health technology
IRB: Institutional Review Board
LMM: Linear mixed-effects model
Q: Question
SD: Standard deviation

## 7 Declarations

### 7.1 Ethics approval and consent to participate

This study adhered to the guidelines outlined in the Declaration of Helsinki – Ethical Principles for Medical Research Involving Human Subjects. The study protocol was approved by the Institutional Review Board (ADVARRA Center for IRB Intelligence (CIRBI)). The ARC Study has been registered at ClinicalTrials.gov as NCT06885918. All participants provided informed consent prior to any study procedures. There was no compensation for participation in the study.

### 7.2 Consent for publication

Not applicable.

### 7.3 Availability of data and materials

Data processing and statistical analyses were completed in MATLAB R2024b (MathWorks, Natick, MA).

### 7.4 Competing interests

The authors declare the following competing interests:

MS has served as a paid consultant for Regeneron. NC – reports no competing interests.

KMB – reports no competing interests. SM - reports no competing interests. HST – reports no competing interests. AP – reports no competing interests.

ALS TDI, under FGVs leadership, has received research support from MT Pharma of America. He has served as a paid scientific reviewer for the ALS CDMRP. He has served as a paid consultant for Guidepoint Global. He acts as an unpaid scientific advisor for ALS Investment Fund.

JDB has received research support from Biogen, MT Pharma of America, MT Pharma Holdings of America, Rapa Therapeutics. He has served as a paid consultant for MT Pharma of America and MT Pharma Holdings of America, Regeneron, Roon, and Alexion. He served as a paid member of a data and safety monitoring board for Sanofi. He acts as an unpaid scientific advisor for the non-profit organizations ALS One and Everything ALS.

### 7.5 Funding

This research has received no funding.

### 7.6 Authors contributions

Concept and design – MS, NC, KMB, SM, HST, AP, FGV, JDB. Data collection – AP, FGV.

Statistical analysis – MS.

Interpretation of results – MS, KMB, NC, JDB. Figure preparation – MS.

Manuscript preparation – MS.

Critical review of manuscript – NC, KMB, SM, HST, AP, FGV, JDB. Study supervision – JDB.

## Notes

### Author Declarations

This study adhered to the guidelines outlined in the Declaration of Helsinki Ethical Principles for Medical Research Involving Human Subjects. The study protocol was approved by the Institutional Review Board (ADVARRA Center for IRB Intelligence (CIRBI)). The ARC Study has been registered at ClinicalTrials.gov as NCT06885918. All participants provided informed consent prior to any study procedures. There was no compensation for participation in the study.

