## Supplementary Materials for "The association between at-home exercise digital metrics and ALS disease progression in lower limbs"

### 1.1 Baseline correlations

When the same exercise metrics were estimated from the right ankle, correlations with ALSFRS-RSE total score remained weak-to-moderate and in the expected direction: count (r = 0.13), duration (r = −0.23), intensity (r = 0.34), and similarity (r = 0.30). The strongest correlations were again observed for the gross motor subdomain (Q7—9), with positive associations for count (r = 0.28), intensity (r = 0.56), and similarity (r = 0.48), and a negative association for duration (r = −0.38). Correlations with bulbar (Q1—3), fine motor (Q4—6), and respiratory (Q10—12) subdomains were smaller, consistent with the lower limb specificity of the task.

A similar pattern was observed when metrics from both ankles were combined. Correlations with ALSFRS-RSE total score were r = 0.20 for count, r = −0.25 for duration, r = 0.34 for intensity, and r = 0.32 for similarity. Associations with gross motor function were stronger, including r = 0.34 for count, r = −0.41 for duration, r = 0.60 for intensity, and r = 0.51 for similarity. These results indicate that baseline associations between exercise-derived metrics and ALSFRS-RSE were reproducible across sensor configurations, with intensity and similarity showing the most consistent alignment with gross motor functioning.

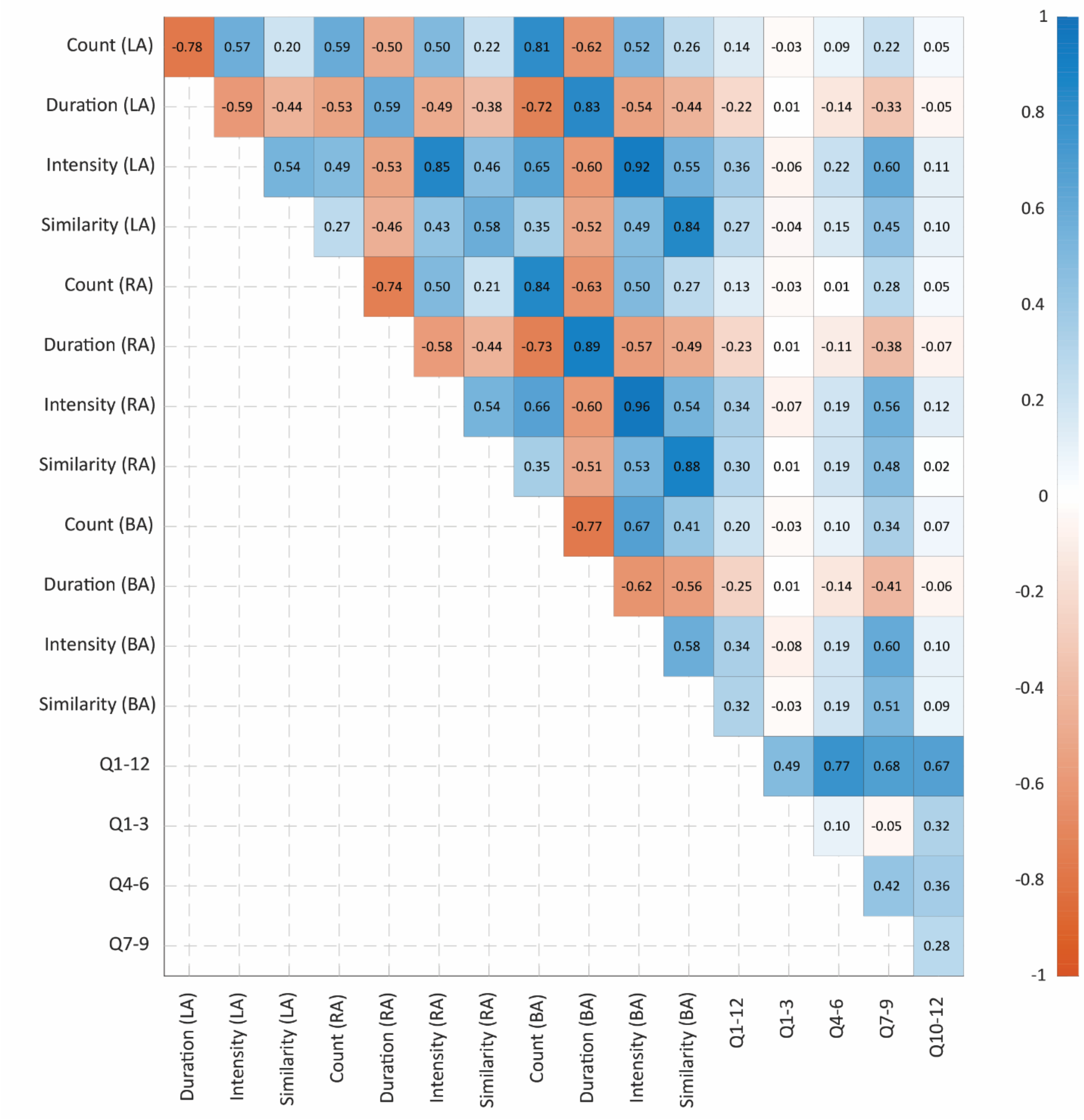


**Figure S1.** Pearson correlation coefficients estimated between baseline digital swing metrics and ALSFRS-RSE total (Q1—12) and subdomain scores (Q1—3 bulbar, Q4—6 fine motor, Q7—9 gross motor, Q10—12 respiratory). Digital metrics were estimated using data collected on the left ankle (LA), right ankle (RA), or both ankles (BA).

### 1.2 Longitudinal change

The sensitivity analysis showed that the longitudinal patterns observed for the left ankle were reproduced when exercise metrics were derived from the right ankle and from both ankles combined. Given that primary findings were not specific to the left ankle sensor location and consistently indicated progressive slowing, reduced vigor, and reduced consistency of repeated lower limb movements across sensor configurations.

**Table S1**. Average baseline and monthly change in ALSFRS-RSE scores and the right and both limb swing movements metrics. Standardized and relative changes are expressed as SD or % per 52 weeks, respectively.

| **Device location** | **Outcome** | **Baseline estimate [95% CI]** | **Monthly change estimate [95% CI]** | ***p*-value** | **R2c** | **R2m** | **Standardized change (SD per year)** | **Relative change (% per year)** |
|---|---|---|---|---|---|---|---|---|
| **Right ankle** | | | | | | | | |
| | **Count** | 43.76 [42.67, 44.84] | -0.017 [-0.095, 0.061] | 0.665 | 0.852 | <0.001 | -0.014 | -0.470 |
| | **Duration** | 1.17 [1.13, 1.20] | 0.007 [0.003, 0.011] | 0.001 | 0.830 | 0.021 | 0.147 | 6.82 |
| | **Intensity** | 0.534 [0.499, 0.568] | -0.006 [-0.008, -0.004] | <0.001 | 0.923 | 0.045 | -0.170 | -12.77 |
| | **Similarity** | 0.847 [0.832, 0.863] | -0.002 [-0.003, -0.002] | <0.001 | 0.840 | 0.034 | -0.168 | -3.51 |
| **Both ankles** | | | | | | | | |
| | **Count** | 41.25 [40.16, 42.34] | -0.075 [-0.151, 0.000] | 0.051 | 0.896 | 0.006 | -0.065 | -2.19 |
| | **Duration** | 1.25 [1.21, 1.29] | 0.011 [0.006, 0.015] | <0.001 | 0.877 | 0.032 | 0.193 | 10.11 |
| | **Intensity** | 0.486 [0.454, 0.518] | -0.006 [-0.008, -0.004] | <0.001 | 0.937 | 0.054 | -0.195 | -14.67 |
| | **Similarity** | 0.802 [0.783, 0.822] | -0.003 [-0.004, -0.002] | <0.001 | 0.859 | 0.037 | -0.176 | -4.61 |

Note: R2c – R-squared conditional, R2m – R-squared marginal.

### 1.3 Digital metrics vs. ALSFRS-RSE

**Table S2.** Number of study participants ($n_s$) along with number of question responses ($n_r$) used to assess the association between digital metrics and ALSFRS-RSE.

| | **Q7** | **Q8** | **Q9** |
|---|---|---|---|
| **Response = 4 ($n_s$, $n_r$)** | 188, 1716 | 136, 1214 | 128, 1139 |
| **Response = 3 ($n_s$, $n_r$)** | 257, 1860 | 189, 1210 | 218, 1374 |
| **Response = 2 ($n_s$, $n_r$)** | 179, 1159 | 225, 2139 | 167, 866 |
| **Response = 1 ($n_s$, $n_r$)** | 66, 280 | 82, 488 | 150, 926 |
| **Response = 0 ($n_s$, $n_r$)** | 21, 56 | 12, 20 | 119, 766 |

### 1.4 Comparison with free-living metrics

Comparably to the main findings, among exercise metrics, count remained stable over time (monthly change = −0.003; 95% CI −0.030, 0.024; $p = 0.818$), while duration increased modestly (0.002; 95% CI 0.000, 0.003; $p = 0.024$) and intensity decreased modestly (−0.001; 95% CI −0.001, −0.000; $p = 0.010$). In contrast, all free-living metrics changed significantly over time, including declining step counts (−17.81 steps/day per month; 95% CI −21.25, −14.37; $p < 0.001$), cadence (−0.284; 95% CI −0.341, −0.228; $p < 0.001$), and stride intensity (−0.003; 95% CI −0.003, −0.002; $p < 0.001$), together with increasing variability (0.001; 95% CI 0.000, 0.001; $p < 0.001$) and fragmentation (0.002; 95% CI 0.001, 0.002; $p < 0.001$). Standardized annual changes were also larger for free-living metrics, ranging from −0.437 SD for cadence to 0.499 SD for fragmentation, compared with −0.106 SD for exercise intensity and 0.189 SD for exercise duration.

**Table S3**. Average baseline and monthly change in exercise and free-living metric estimated from left ankle sensor data.

| | **Outcome** | **Baseline estimate [95% CI]** | **Monthly change estimate [95% CI]** | ***p*-value** | **R2c** | **R2m** | **Standardized change (SD per year)** | **Relative change (% per year)** |
|---|---|---|---|---|---|---|---|---|
| **Exercise** | | | | | | | | |
| | **Count** | 45.26 [43.89, 46.64] | -0.003 [-0.030, 0.024] | 0.818 | 0.844 | <0.001 | -0.011 | -0.37 |
| | **Duration** | 1.14 [1.10, 1.18] | 0.002 [0.000, 0.003] | 0.024 | 0.922 | 0.016 | 0.189 | 8.25 |
| | **Intensity** | 0.557 [0.517, 0.598] | -0.001 [-0.001, -0.000] | 0.010 | 0.898 | 0.016 | -0.106 | -7.68 |

| | | | | | | | | |
|---|---|---|---|---|---|---|---|---|
| | **Similarity** | 0.852 [0.832, 0.871] | -0.000 [-0.001, 0.000] | 0.157 | 0.781 | 0.006 | -0.069 | -1.41 |
| **Free-living** | | | | | | | | |
| | **Step counts** | 3945 [3652, 4239] | -17.81 [-21.25, -14.37] | <0.001 | 0.807 | 0.170 | -0.339 | -23.47 |
| | **Cadence** | 76.29 [72.85, 79.72] | -0.284 [-0.341, -0.228] | <0.001 | 0.829 | 0.202 | -0.437 | -19.39 |
| | **Intensity** | 0.977 [0.936, 1.02] | -0.003 [-0.003, -0.002] | <0.001 | 0.863 | 0.186 | -0.394 | -15.46 |
| | **Similarity** | 0.570 [0.552, 0.588] | -0.000 [-0.001, -0.000] | <0.001 | 0.714 | 0.031 | -0.148 | -4.49 |
| | **Variability** | 0.419 [0.406, 0.433] | 0.001 [0.000, 0.001] | <0.001 | 0.851 | 0.076 | 0.265 | 8.46 |
| | **Fragmentation** | 0.476 [0.457, 0.494] | 0.002 [0.001, 0.002] | <0.001 | 0.862 | 0.184 | 0.499 | 20.50 |

Note: R2c – R-squared conditional, R2m – R-squared marginal.

### 1.5 Onset impact

For right ankle exercise metrics (**Figure S2**), the pattern was directionally consistent with the left ankle analysis: participants with right lower limb onset had lower baseline count, higher duration, lower intensity, and lower similarity compared with participants without right lower limb onset. The difference in count did not reach statistical significance ($p$ = 0.0608), whereas duration was significantly higher in the onset group ($p$ = 0.0073). Intensity and similarity were also significantly lower among participants with right lower limb onset, with $p$-values of 9.86e-09 and 7.46e-06, respectively. Thus, as in the left ankle analysis, exercise quality metrics were more strongly differentiated by reported side of onset than repetition count.

When exercise metrics were calculated using data from both ankles (**Figure S3**), the associations with lower limb onset were attenuated. Participants with onset on either lower limb again tended to show lower count, longer duration, lower intensity, and lower similarity compared with participants without lower limb onset, but only intensity reached statistical significance ($p$ = 0.0042). Count ($p$ = 0.3817), duration ($p$ = 0.0947), and similarity ($p$ = 0.0671) did not differ significantly between groups. These results suggest that side-specific sensor placement may be more sensitive to the anatomical site of ALS onset than bilateral aggregation, particularly for detecting reduced movement intensity and consistency in the affected lower limb.

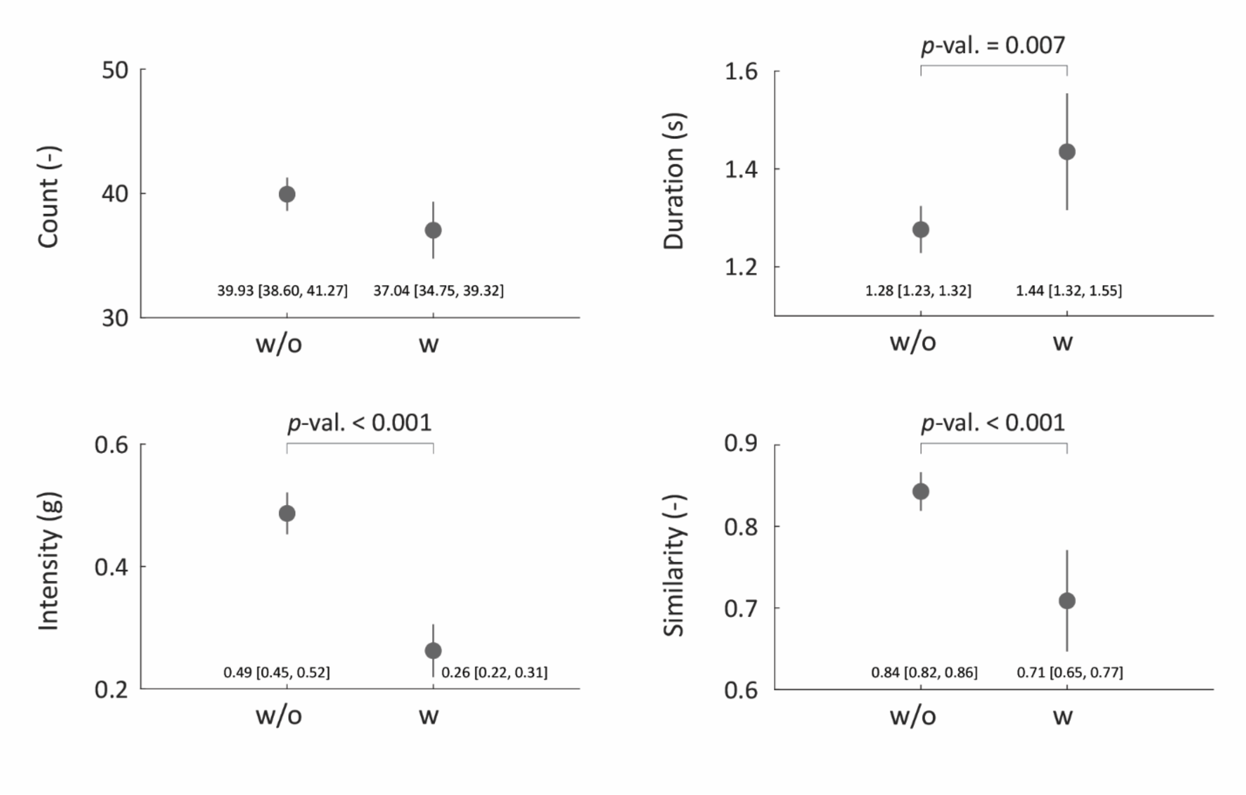


**Figure S2.** Mean and 95% confidence intervals for baseline digital metrics (first valid set of observations) stratified on participants with (w) and without (w/o) ALS onset on right leg.

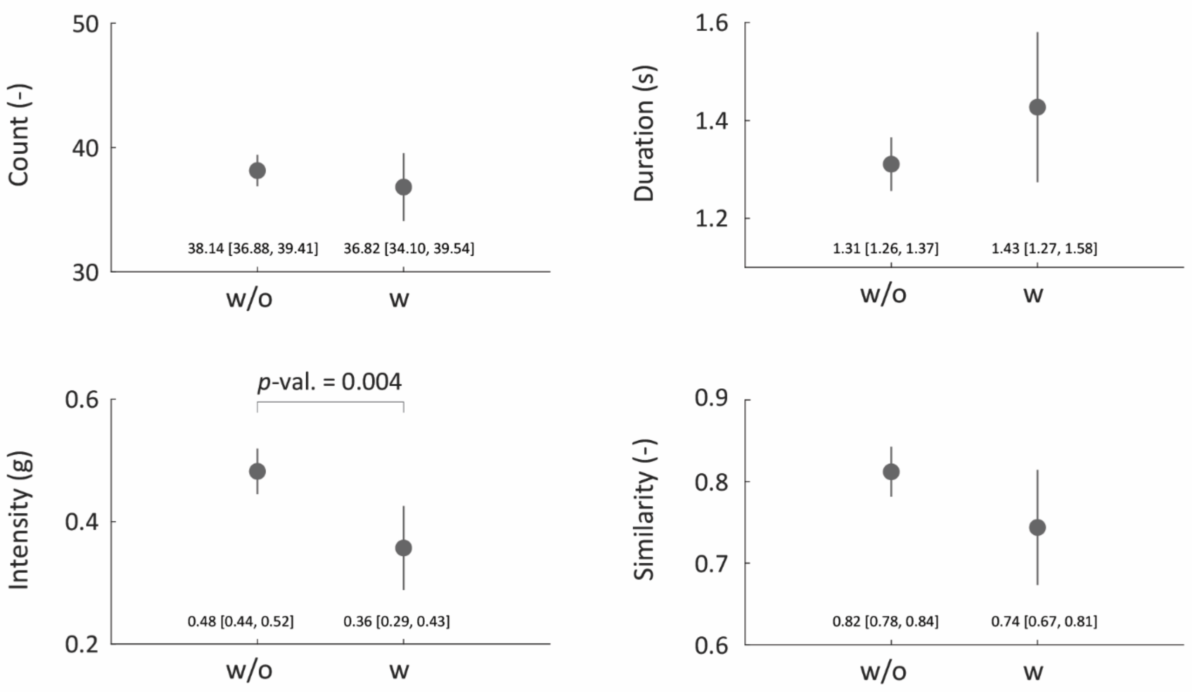


**Figure S3**. Mean and 95% confidence intervals for baseline digital metrics (first valid set of observations) stratified on participants with (w) and without (w/o) ALS onset on either side. Here exercise metrics were calculated using data from both sensors.

### 1.6 Normal vs. deteriorated lower limb functioning at baseline

For the right ankle, baseline values were again higher in the normal-functioning group for count (48.10; 95% CI 46.32, 49.87 vs. 42.02; 95% CI 40.74, 43.3), intensity (0.7776; 95% CI 0.724, 0.831 vs. 0.435; 95% CI 0.399, 0.471), and similarity (0.918; 95% CI 0.906, 0.931 vs. 0.818; 95% CI 0.798, 0.839), and lower for duration (1.02; 95% CI 0.98, 1.05 vs. 1.23; 95% CI 1.18, 1.27). In longitudinal models, the deteriorated group showed significant worsening in duration (0.010; 95% CI 0.005, 0.015; $p < 0.001$), intensity (−0.007; 95% CI −0.009, −0.005; $p < 0.001$), and similarity (−0.003; 95% CI −0.004, −0.002; $p < 0.001$), while count did not reach statistical significance ($p = 0.106$). Slope differences between groups were significant for count ($p = 0.006$) and duration ($p = 0.011$), but not for intensity ($p = 0.168$) or similarity ($p = 0.113$).

When metrics were computed using both ankles, results were consistent with the single-ankle analyses. The deteriorated group had lower baseline count (39.11; 95% CI 37.85, 40.37 vs. 46.59; 95% CI 44.88, 48.30), higher duration (1.33; 95% CI 1.28, 1.38 vs. 1.05; 95% CI 1.01, 1.09), lower intensity (0.387; 95% CI 0.355, 0.420 vs. 0.730; 95% CI 0.681, 0.780), and lower similarity (0.763; 95% CI 0.738, 0.788 vs. 0.900; 95% CI 0.885, 0.916). In the deteriorated group, all four bilateral metrics changed significantly over time, with decreasing count (−0.144; $p = 0.003$), increasing duration (0.015; $p < 0.001$), decreasing intensity (−0.006; $p < 0.001$), and decreasing similarity (−0.004; $p < 0.001$). As with the right ankle analysis, slope differences were significant for count ($p = 0.003$) and duration ($p = 0.004$), but not for intensity ($p = 0.394$) or similarity ($p = 0.112$), supporting the robustness of the stratified findings across sensor configurations**.**

The results for the right ankle and both ankles are summarized in **Table S3**.

**Table S4.** Comparison between longitudinal changes in exercise metrics collected from the right ankle and a combination of both ankles between individuals reporting normal and deteriorated functioning, as reflected on questions Q8 and Q9 in ALSFRS-RSE. Model slopes express monthly change in digital metrics and survey scores. Standardized and relative changes are expressed in SD and percentage change per year, respectively.

| **Device location** | **Digital metric** | **Intercept** | **Slope** | ***p*-value** | **R2c** | **R2m** | **Standardized change (SD per year)** | **Relative change (% per year)** | **Intercept** | **Slope** | ***p*-value** | **R2c** | **R2m** | **Standardized change (SD per year)** | **Relative change (% per year)** | **Slope difference p-value** |
|---|---|---|---|---|---|---|---|---|---|---|---|---|---|---|---|---|
| | | **Normal at baseline, n=100** | | | | | | | **Deteriorated at baseline, n=249** | | | | | | | |
| **Right ankle** | | | | | | | | | | | | | | | | |
| | **Count** | 48.10 [46.32, 49.87] | 0.141 [0.047, 0.236] | **0.003** | 0.745 | 0.030 | 0.124 | 3.52 | 42.02 [40.74, 43.3] | -0.083 [-0.183, 0.018] | 0.106 | 0.868 | 0.007 | -0.071 | -2.37 | **0.006** |
| | **Duration** | 1.02 [0.98, 1.05] | -0.002 [-0.004, 0.001] | 0.124 | 0.786 | 0.012 | -0.097 | -2.24 | 1.23 [1.18, 1.27] | 0.010 [0.005, 0.015] | **<0.001** | 0.837 | 0.037 | 0.199 | 9.83 | **0.011** |
| | **Intensity** | 0.7776 [0.7238, 0.8314] | -0.004 [-0.008, 0.001] | 0.149 | 0.864 | 0.013 | -0.105 | -5.41 | 0.435 [0.399, 0.471] | -0.007 [-0.009, -0.005] | **<0.001** | 0.926 | 0.085 | -0.228 | -18.01 | 0.168 |
| | **Similarity** | 0.9183 [0.9055, 0.9311] | -0.001 [-0.002, -0.000] | 0.043 | 0.625 | 0.015 | -0.118] | -1.34 | 0.818 [0.798, 0.839] | -0.003 [-0.004, -0.002] | **<0.001** | 0.855 | 0.041 | -0.182 | -4.28 | 0.113 |
| **Both ankles** | | | | | | | | | | | | | | | | |
| | **Count** | 46.59 [44.88, 48.30] | 0.097 [-0.000, 0.193] | 0.050 | 0.804 | 0.015 | 0.088 | 2.49 | 39.11 [37.85, 40.37] | -0.144 [-0.240, -0.049] | **0.003** | 0.905 | 0.022 | -0.131 | -4.43 | **0.003** |

| | | | | | | | | | | | | | | | |
|---|---|---|---|---|---|---|---|---|---|---|---|---|---|---|---|
| | **Duration** | 1.05 [1.01, 1.09] | -0.001 [-0.004, 0.002] | 0.465 | 0.840 | 0.003 | -0.056 | -1.33 | 1.33 [1.28, 1.38] | 0.015 [0.009, 0.022] | **<0.001** | 0.882 | 0.052 | 0.251 | 13.76 | **0.004** |
| | **Intensity** | 0.730 [0.681, 0.780] | -0.005 | 0.050 | 0.878 | 0.023 | -0.149 | -7.57 | 0.387 [0.355, 0.420] | -0.006 [-0.008, -0.005] | **<0.001** | 0.945 | 0.095 | -0.258 | -19.92 | 0.394 |
| | **Similarity** | 0.900 [0.885, 0.916] | -0.001 | 0.022 | 0.630 | 0.020 | -0.131 | -1.80 | 0.763 [0.738, 0.788] | -0.004 [-0.005, -0.002] | **<0.001** | 0.871 | 0.046 | -0.192 | -5.71 | 0.112 |

Note: R2c – R-squared conditional, R2m – R-squared marginal.
